# A Multicenter Retrospective Observational Cohort Study of Seizure Freedom before Lennox-Gastaut Syndrome (the “Gap”). Opportunities for Prevention

**DOI:** 10.1101/2024.12.03.24318373

**Authors:** Laura Deering, Aaron Nelson, Elissa Yozawitz, Steven Wolf, Patricia McGoldrick, Alan Wu, Natasha Basma, Zachary Grinspan

## Abstract

**Objective:** Lennox-Gastaut Syndrome (LGS) is a severe, often treatment-resistant epilepsy syndrome typically diagnosed in early childhood. Many have seizures before diagnosis. Some have periods of seizure freedom before treatment resistance, i.e., a “gap.” Review of these gaps may identify early candidate biomarkers of LGS and/or highlight opportunities for intervention.

**Methods:** We reviewed charts of children diagnosed with LGS born in 2008-2010 and diagnosed with LGS by 2014 at five academic medical centers in New York City using the RENYC (Rare Epilepsies in New York City) database. We collected dates of events of potential biomarkers by chart abstraction, including onset of slow spike-and-wave (SSW) and onset and offset of seizure freedom. Seizure-free periods (“gaps”) were defined as greater than 30 days without unprovoked seizures.

**Results:** Thirty-three children had LGS (52% male; etiology 33% structural-acquired, 6% structural-congenital, 3% genetic-structural, 24% genetic, 33% unknown). Twenty-two (67%) had a gap before diagnosis. Eight of these twenty-two (36%) had SSW described before the gap, five (23%) during the gap, and six (27%) after the gap. A history of infantile epileptic spasms syndrome (IESS), age at seizure onset, and age of tonic seizure onset were not different between those with and without a gap. Of 20 (61%) with a history of IESS, 10 (30% of the full cohort) had not received recommended therapy (i.e., ACTH, prednisolone, or vigabatrin) as first-line treatment.

**Conclusions:** The appearance of SSW, even in seizure-free children, may herald the development of LGS in high-risk children. Further studies on its predictive value are warranted. Our findings also highlight use of recommended first-line therapy for infantile spasms as a potentially modifiable treatment gap in children who subsequently develop LGS.

## INTRODUCTION

Lennox-Gastaut syndrome (LGS) is a severe, often treatment-resistant form of epilepsy characterized by the triad of multiple seizure types, cognitive impairment, and a slow spike-and-wave (SSW) pattern on EEG. These features frequently emerge separately over time. Children typically meet criteria before age of eight years, most commonly between age three and five. LGS often persists into adulthood with treatment-resistant seizures, ongoing cognitive and physical disabilities,^1,2^ and higher mortality than the general population.^3,4^

LGS may be due to genetic, acquired, or unknown etiologies.^5,6^ Most commonly, regardless of etiology, seizures begin before the LGS diagnosis.^4,7,8^ However, the progression to LGS is under-described. For some, epilepsy begins as a developmental and epileptic encephalopathy, like infantile epileptic spasms syndrome, whereas for others, epilepsy may initially be non-syndromic. For many, there is a seizure-free gap between when early-life epilepsy resolves and LGS begins.^4^ It is unknown if there are early biomarkers that can identify which children will evolve to LGS. Identifying early candidate biomarkers could be valuable, as there may be interventions to prevent progression to LGS.

One potential set of biomarkers heralding future LGS are EEG changes. Clinical experience suggests that SSW sometimes emerges before seizure recurrence. Some evidence suggests the mean age at first SSW in a population of LGS is between 3 and 6 years.^9,10^ However, the relationship between SSW, seizure control, and LGS onset is under-described.

Clarifying when features of LGS first occur might provide an opportunity for earlier intervention to prevent progression to LGS. There are already examples of such interventions. Surgical removal of epileptogenic lesions can lead to complete resolution of LGS,^11–13^ and earlier epilepsy surgery is associated with better outcomes.^14,15^ In addition, hormonal treatment can lead to permanent remission of epileptic encephalopathies such as IESS and electrographic status epilepticus of sleep^16,17^ – though it is unclear if it can prevent LGS. The current work aims to identify opportunities for intervention to prevent progression to encephalopathy by examining the evolution of LGS over the first 4 to 6 years of life.

## METHODS

### Study Design

We performed a retrospective cohort study of children with LGS using the Rare Epilepsies in New York City (RENYC) database. RENYC includes neurology notes written between 2010 – 2014 from five academic medical centers in New York City: Weill Cornell Medicine, Columbia University Medical Center, New York University Langone Medical Center, Montefiore Medical Center, and Mount Sinai Health System. The institutional review board at all five institutions reviewed and approved this study, facilitated by a central IRB (Biomedical Research Alliance of New York – BRANY).

### Subjects

Candidate cases of LGS were flagged for review if the notes included key words associated with LGS.^18^ Three reviewers (ZG, NB, and DH) examined the flagged cases and made an initial determination if the child had LGS. The diagnosis was confirmed by an in-depth chart review by a fourth reviewer (LD). One author (ZG) provided the final adjudication if there were discrepancies or questions. Reviewers examined cases at their own institution and at other institutions. Sometimes, a reviewer had previously provided for a person in the study.

### Definition of LGS

A diagnosis of LGS required all three of (1) developmental delay, (2) multiple seizure types, and (3) description of SSW on an EEG report. We selected this working definition of LGS after reviewing published case definitions from several sources.^4,8,19–22^ We did not require tonic seizures, as there is uncertainty when these begin. To focus on the early evolution of LGS, we examined children who turned 4, 5, or 6 in 2014 (i.e., those with 4-5 years of detailed medical records starting in the first year of life).

### Inclusion/Exclusion Criteria

We required children to have at least two healthcare encounters per year and clinical notes describing seizure frequency, seizure type, and important events. We also required description of SSW on at least one EEG report.

### Chart Abstraction Process

After reviewing a subset of charts, we created, iterated, and finalized a taxonomy of important events in the natural history of LGS. These events included: date of birth, first seizure, first and last visits in the RENYC database, first (if any) instance of hypsarrhythmia in the EEG reports, first instance of SSW in the EEG, first tonic seizure, any instance of status epilepticus, any epilepsy monitoring unit (EMU) visits, epilepsy-related hospitalizations, and onset and offset of seizure freedom. We then reviewed the full cohort to catalog the date of each event. If an event happened in a certain month but the day was not specified, we used the 15^th^ of the month. Hypsarrhythmia and SSW were identified based on the EEG reports (the raw EEG tracings were unavailable). A description of spike-and-wave discharges at 2.5 Hz or lower was considered “slow”.

We used notes from inpatient and outpatient encounters to reconstruct seizure history. Seizure-free periods (i.e., gaps) were defined as greater than 30 days without unprovoked seizures. We defined the length of each gap from the first of those 30 until the day seizures recurred. If the exact time of the last seizure was unknown, but a clinician wrote a note that the patient was seizure-free, the visit date was considered the start date of the gap. If the patient was seizure-free at the last visit, the end date of the gap was the date of the last visit.

During the chart abstraction process, we also noted the following: any diagnosis of IESS, ever use of a gastrostomy tube (G-tube) or ventriculoperitoneal (VP) shunt, ever epilepsy surgery. If there was a history of IESS, we noted age at spasm onset, presence of hypsarrhythmia with spasm onset, and time from clinical onset to first-line treatment. We also noted if the follow-up EEG showed epileptiform discharges and if hypsarrhythmia resolved within two weeks of treatment. If epileptic spasms resolved within three months of diagnosis, we noted the number of anti-seizure medications before resolution, if there was a relapse of epileptic spasms, when the relapse occurred, and how it was treated. Finally, we noted if children had other seizure types by age one.

We grouped LGS etiology into mutually exclusive categories, as per Wirrell et al^23^, in rough alignment with the International League Against Epilepsy etiology framework.^24^ They are: genetic, genetic-structural, structural-congenital, structural-acquired, metabolic, immune, infectious, and unknown. We reviewed the MRI reports for those with structural-congenital, structural-acquired, or unknown etiologies. We noted cortical injury or abnormality (yes/no), extent of white matter injury (none, scattered, patchy, extensive), presence of deep grey or brainstem abnormality (yes/no), and state of the corpus callosum (normal, thin, partially missing, absent).

### Visual Analysis

We created a visual vocabulary for the clinical trajectory of the subjects from birth, guided by a hierarchy of visual perception described by Cleveland and McGill.^25^ For example, events are positioned along a common scale to allow examination of each individual’s history and comparison across different patients’ histories. Length shows gaps and allows for visualization within and across patients. The color and shape of symbols highlight events of interest. To avoid visual clutter, the visualization includes only the first seizure, first tonic seizure, first visit in the RENYC database, last visit, and gap.

### Statistical Analysis & Cumulative Incidence Plots

We used median statistics (median and interquartile interval (IQI)) for univariate descriptions and bivariate comparisons. When distributions were very skewed, we calculated both IQI and absolute range. We compared characteristics of children with vs. without a gap, using chi-square tests for categorical variables and Mann-Whitney U test for continuous variables. If bivariate analyses had been significant at *p* < 0.05, we had planned multivariable logistic regressions. We highlight effect sizes that appeared clinically important, even if not statistically significant. We provide two cumulative incidence plots to illustrate the timeframe of development of clinical features. All statistics and visualizations were conducted in R, version 4.0.2 (R Foundation for Statistical Computing).

### Clinician Review

The authors had robust discussions about the data, including speculations about the implications of the data and the next steps. We include a summary of these meetings in the discussion section.

## RESULTS

There were 75 children with suspected LGS aged 4–6 years in 2014 based on the initial review process. After review, we excluded 42, for the following reasons. Nine did not meet all diagnostic criteria for LGS (4 without development delay, 2 with refractory epileptic spasms that had not progressed to LGS, 1 with Doose syndrome, and 2 with no EEG report indicating SSW). Twenty-six had only one note. Seven had insufficient information about seizure frequency.

The final cohort included 33 children -- 6 born in 2008, 17 in 2009, and 10 in 2010. The etiology was unknown for 11 (33%) children, including children for whom genetic testing was limited or inconclusive. Eleven (33%) had a structural-acquired etiology (e.g., hypoxic-ischemic encephalopathy, trauma, or infection). Eight (24%) had known genetic mutations including CDKL5, KCNT1, SCN2A, Trisomy 21, Snyder Robinson Syndrome (SMS mutation), duplication of the short arm of the X chromosome (p11.22-11.223), and a supernumerary ring chromosome 4. Two (6%) had congenital structural brain malformations (e.g., Aicardi syndrome) and one (3%) had a known genetic mutation causing a structural brain malformation (Lis1). (Table 1)

**Table 1.**
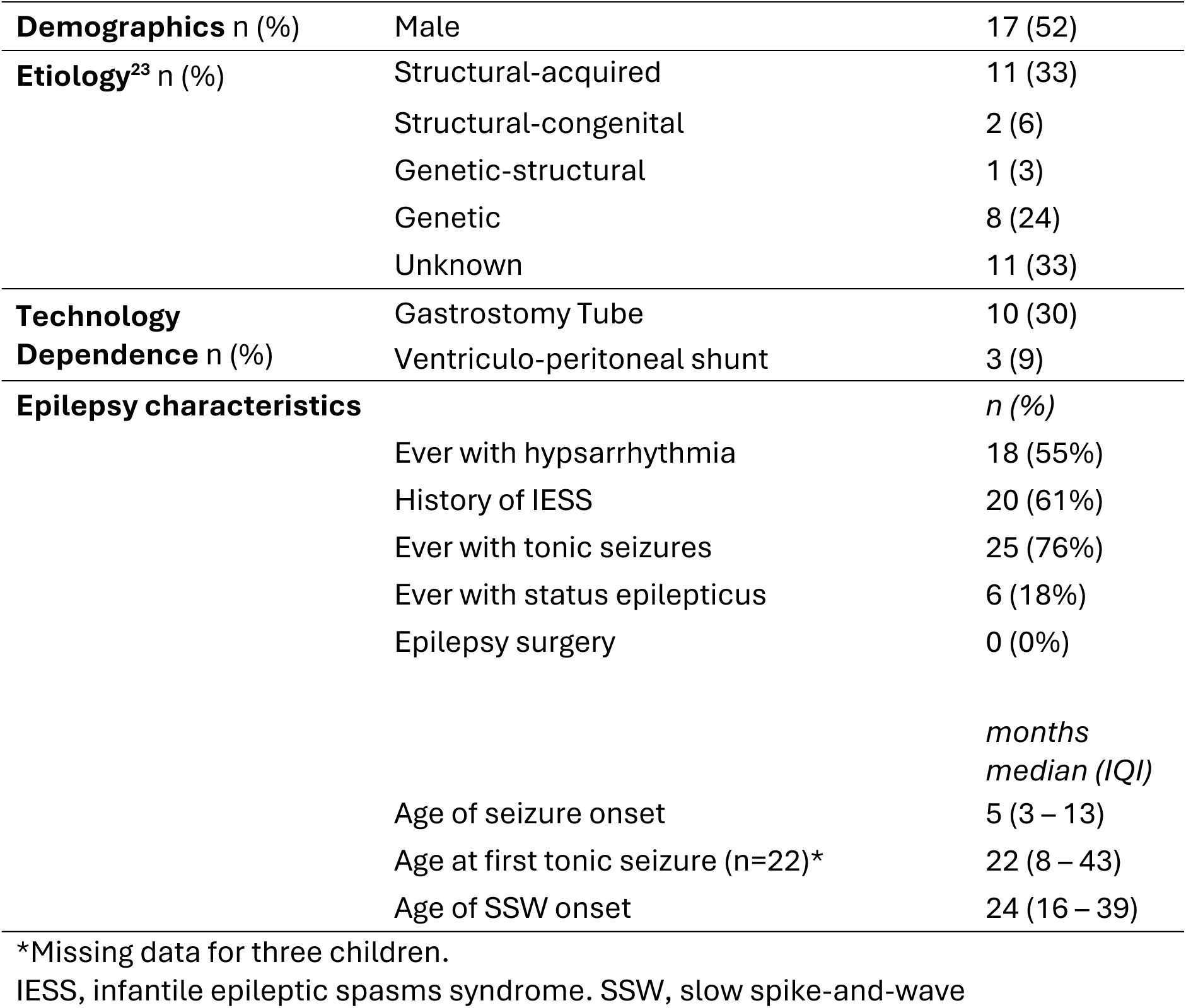
Patient characteristics of 33 children with LGS born 2008 - 2010.

For the 11 with unknown etiologies, one had no MRI reported and 10 had normal MRIs. For the two patients with congenital structural abnormalities, one had lissencephaly, and the other had a cortical injury, extensive white matter changes, and absent corpus callosum.

For the 11 with acquired structural etiologies, seven had cortical injuries and eight had white matter injuries, all extensive. One had brainstem injury and four had corpus callosum abnormalities.

The median onset of seizures was 5 months (IQI 3 – 13); the median onset of SSW wave was 24 months (IQI 16 – 39). Twenty (61%) had IESS before LGS and 18 (55%) had hypsarrhythmia on at least one EEG report. Twenty-five patients (76%) had tonic seizures. For 3 of these 25, the onset of tonic seizures was uncertain – though they were present by 12 months, 14 months, and 3 years. For the remaining 22, the median age at onset was 22 months (IQI 8 – 43). Interestingly, among 22 children with a known date of onset of tonic seizures, there were 7 (32%) where the tonic seizures occurred *before* the first report of SSW. Six children (18%) had at least one episode of status epilepticus. Ten (30%) were G-tube dependent at some point during their course. Three (9%) had a VP shunt. None had epilepsy surgery. (Figure 2 & 3)

**Figure 1.**
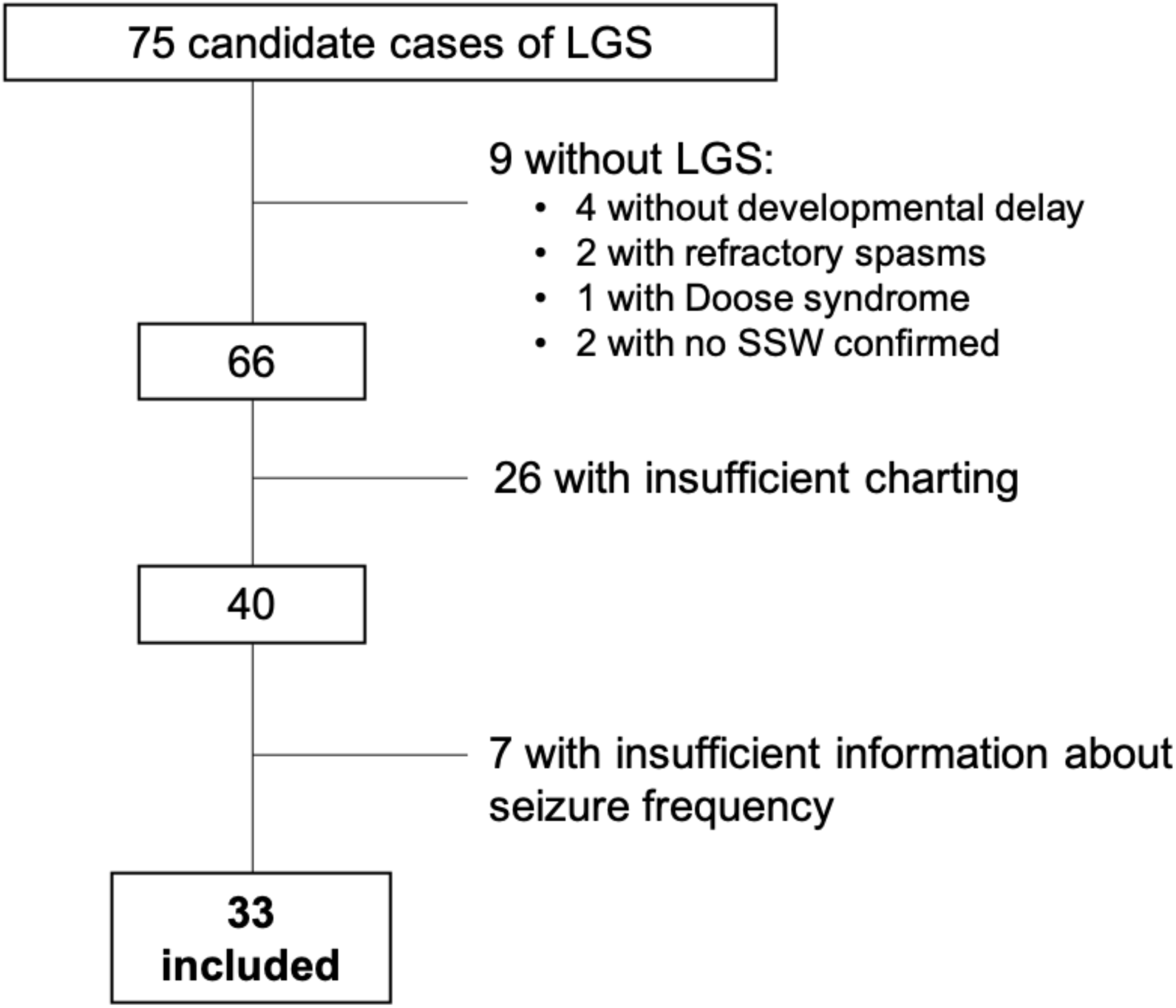
Children included and excluded from the study. SSW = Slow spike and wave

**Figure 2.**
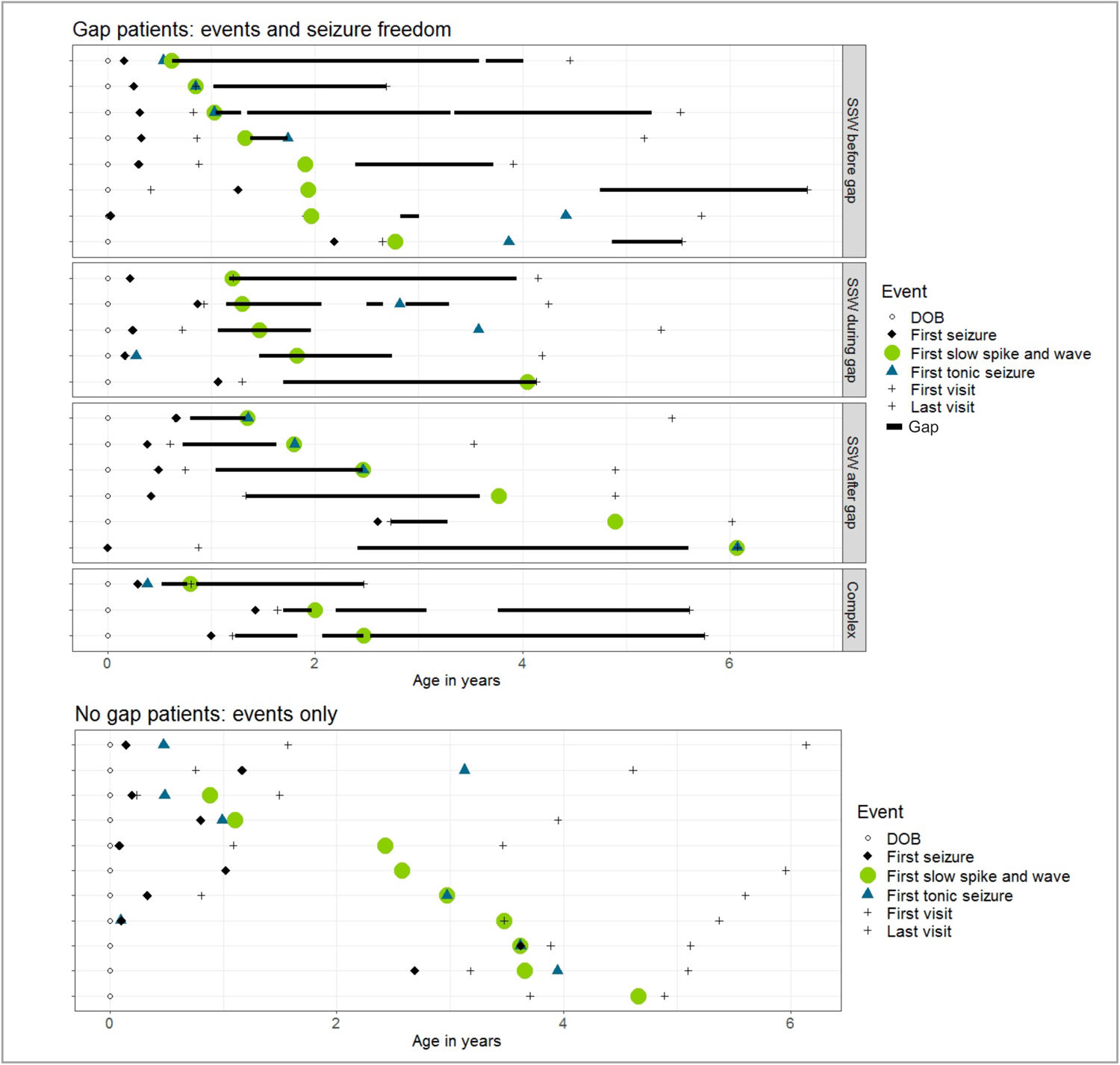
Visualization of the important events captured for patients with a gap (top) and patients without a gap (bottom). Each row is a unique patient. In the top image, patients are grouped based on timing of the slow spike-and-wave in relation to the gap. Gray bars in the same image represent the length of the gap. DOB = date of birth; EMU = epilepsy monitoring unit.

**Figure 3.**
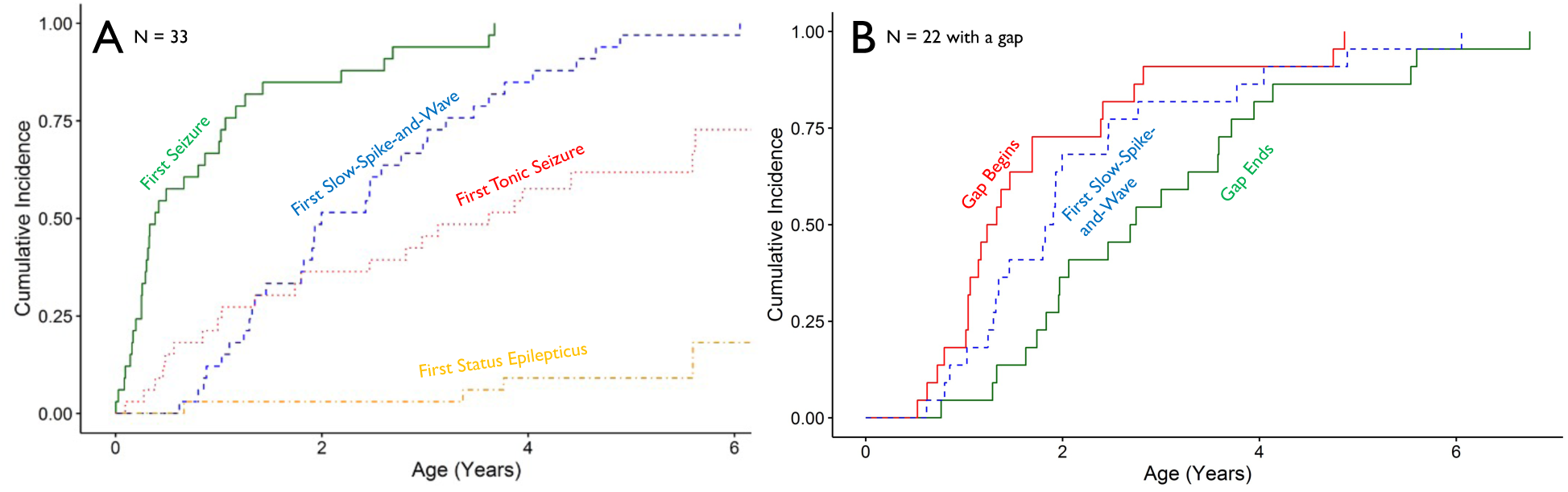
Cumulative incidence plots of early events in the evolution of Lennox-Gastaut Syndrome for (A) 33 children in the cohort and (B) a subset of 22 children with a gap. (A) Green solid line – first seizure; red dotted line – first tonic seizure; blue dashed line – first slow spike-and-wave noted on EEG; orange dot-dash – first episode of status epilepticus. (B) Red solid line – time when gap begins; blue dashed line – first slow spike-and-wave noted on EEG; green solid line – time when gap ends. In (B), the timing of the first slow spike-and-wave (dashed line) occurs during the same period as the gap.

Twenty children (61%) had a prior diagnosis of IESS, starting at a median of 5.5 [IQI 7.9] months; 18 (90%) with hypsarrhythmia. Ten (50%) received recommended first line therapy (ACTH, prednisone, or vigabatrin) which was started, on average, at a median of 3 months after the seizures began [IQI 2.5 months, absolute range less than a month to one year].

Other initial treatments included levetiracetam, phenobarbital, topiramate, valproic acid, zonisamide, and clonazepam. Spasms resolved for 8 (40%) within three months (ongoing spasms for 9, unknown for 3). Of these 8, 4 resolved after 1 medication, 4 required 2 or more. Four had a relapse of spasms on average 3 [range of 2-7] months after initial resolution (for one, the time of relapse was unknown). The relapses were treated ACTH (1), valproic acid (1), vigabatrin (1), or an increased dose of topiramate (1). (Table 2)

**Table 2.** Characteristics of the 20 children with infantile spasms.

|  | Yes<br>n (%) | No<br>n (%) | Unknown<br>n (%) |
| --- | --- | --- | --- |
| Hypsarrhythmia with onset of IESS | 11 (61) | 3<sup>†</sup> (17) | 4 (22) |
| Hypsarrhythmia resolved within 2 weeks of treatment | 4 (24) | 7 (41) | 6 (35) |
| Epileptiform discharges present after treatment | 13 (65) | 2 (10) | 5 (25) |
| Spasms resolved within 3 months of treatment | 8 (40) | 9 (45) | 3 (15) |
| Other seizure types before 1 year of age | 11 (55) | 8* (40) | 0 |

IESS, infantile epileptic spasms syndrome
<sup>†</sup>Two patients did not ever develop hypsarrhythmia, one developed it after treatment began
\*One began having seizures after 1 year of age

Twenty-two (67%) had a period of seizure freedom (a “gap”) greater than 30 days. For 8 (36%), SSW appeared before the gap, 5 (23%) during the gap, and 6 (27%) after the gap. Three (14%) had a complex relationship between their first SSW and seizure frequency – these children had multiple gaps and the SSW appeared between them. (Figure 2) Visual inspection of the cumulative incidence plots demonstrated that the onset of SSW occurred roughly in the same time frame as the first gap (Figure 4).

There were 33 gaps in 22 children with a median length of 11 months (IQI 5 – 23). All children with a gap were undergoing treatment for seizures during the gap. In 17 (52%) of these 33 gaps, children were taking one anti-seizure medication (ASM), and in 6 (18%), two ASMs. For the remaining 10 gaps (30%), ASMs were added or subtracted during the gap (8), or the children were only prescribed hormonal therapy (1 ACTH and 1 prednisolone).

A history of IESS was not associated with a gap -- 14 of 20 children with prior IESS had a gap (70%) vs 8 of 13 without prior IESS (61%) (p = 0.9). Tonic seizures were not statistically more common among children who did not have a gap (10 had tonic seizures among 11 without a gap (91%) vs 15 among 22 with a gap (68%); p = 0.3); however, the 23-percentage point difference may be clinically meaningful. The age of onset of tonic seizures did not differ between children with or without a gap (22 [11 – 44] months with a gap vs 24 [6 – 40] months without, *p* = 0.7; 1 missing). (Table 3)

**Table 3.** Bivariate analyses comparing patients with and without a gap.

| **Continuous variables†** | **Gap (n = 22)** | **No Gap (n = 11)** | **p** |
| --- | --- | --- | --- |
| Age at seizure onset | 4 (3 - 12) | 10 (2 - 24) | 0.55 |
| Age at SSW onset | 23 (16 - 30) | 37 (30 - 43) | 0.07 |
| Age at tonic seizure onset* | 22 (11 - 44) | 24 (6 - 40) | 0.66 |

† Median (interquartile interval) months
\* 3 patients with known tonic seizures not no clear date of onset excluded
SSW, slow spike-and-wave

| **Categorical variables** | **Gap** | **No Gap** | **p** |
| --- | --- | --- | --- |
| History of IESS | 14 of 20 (64%) | 6 of 20 (55%) | 0.90 |
| Tonic seizures | 15 of 25 (68%) | 10 of 25 (91%) | 0.31 |

IESS, infantile epileptic spasms syndrome.

Children with a gap often had key events that occurred earlier in life, though the differences were not statistically significant. They were younger at the onset of their seizures (median 4 [3 – 12] months vs 10 [2 – 24] months; p = 0.55), and younger when the SSW was first described (median 23 [16 – 30] months vs 37 [30 – 43] months; p = 0.07). (Table 3)

## DISCUSSION

### Summary of Findings

Most children with LGS (two-thirds) had early-life seizures followed by a period of seizure freedom (a gap) before diagnosis with LGS. For those with a gap, SSW first appeared before or during the gap for more than half. In addition, those with a gap often had earlier seizure onset or earlier appearance of SSW and were more likely to have tonic seizures by the end of the studied period, though these differences were not statistically significant. Other markers did not differ between children with vs without a gap.

Most children with LGS (61%) had a prior history of IESS. Only half of this subgroup received recommended^26–29^ first-line therapy (i.e., hormonal therapy or vigabatrin). For two-thirds of this subgroup, infantile spasms syndrome was challenging to treat – either refractory to treatment or with a relapse after initial resolution. These data suggest two opportunities for intervention: (1) use of SSW as a marker for more aggressive therapy and (2) improved adherence to treatment recommendations for infantile spasms.

### Context in Literature

These results expand on similar findings by Berg et al.^4^ who also observed periods of seizure freedom in children with LGS, though a smaller proportion (32% in their work vs 67% here). This difference may be due to study design. Their report focuses on year-to-year seizure frequency rather than our more granular view. They drew on parental reporting of seizure frequency over the phone in addition to review of medical records while our study was limited to only medical record review. Their study was a prospective study of incident early-life epilepsy, whereas our study was retrospective of children with prevalent LGS.

### Clinician data review

During a discussion of the results among the clinician authors (AN, ZG, EY, SW, PM), three themes emerged. First, our findings had face validity: we recalled visits from patients with good clinical seizure control despite EEG with epileptiform abnormalities. Second, we did not have good evidence or clinical guidance on how to manage such individuals. Third, we were enthusiastic about a trial in which at-risk patients (e.g., those with a history of IESS or neonatal seizures) were treated prophylactically when an EEG showed SSW during a period of seizure freedom. There was no clear consensus on what that intervention would be. Possibilities discussed included hormonal therapy (corticosteroids or ACTH), vigabatrin, zonisamide, topiramate, levetiracetam, valproic acid, or clobazam.

### Slow spike-and-wave as a marker for more aggressive therapy?

The idea to guide therapy based on EEG findings has precedent – EEG findings guide treatment for IESS^30^ and electrical status epilepticus of sleep.^17^ Furthermore, two prospective trials found that the prophylactic treatment of an abnormal EEG in infants with tuberous sclerosis complex can prevent IESS.^31,32^ For LGS, vigabatrin and hormonal therapies are reasonable options, given the effectiveness of both medications in infantile spasms,^33^ the effectiveness of vigabatrin in tuberous sclerosis,^31,32^ and case series showing improvement of LGS with steroids.^19,34,35^

### Is slow spike-and-wave sufficiently reliable as a biomarker?

The inter-rater reliability to detect SSW on EEG is unknown and needs study. The experience with inter-rater reliability of hypsarrhythmia suggests both caution and optimism -- caution because inter-rater reliability to detect hypsarrhythmia based on unguided physician review is poor;^36^ optimism because inter-rater reliability markedly improved when clinicians were provided a scoring system to identify hypsarrhythmia.^37^ Of additional importance, SSW is not pathognomonic for LGS,^7,8,20^ and can also be seen in other clinical contexts, such as focal epilepsy with secondary bilateral synchrony.

### Recommended Care for Infantile Spasms

Although consensus statements^26,27^ and quality measurement sets^29^ consistently recommend ACTH, oral steroids, or vigabatrin as first-line therapy for IESS, half the children with IESS in our cohort had received a different agent as first-line therapy. Quality improvement initiatives to consistently provide recommended first-line therapy for IESS are warranted and can improve response rates.^38^ The potential effect on subsequent incidence of LGS merits study.

### Tonic seizures as a criterion for LGS

We were surprised that some children diagnosed with LGS did not have tonic seizures. It is unclear if tonic seizures should be conceptualized as a commonly observed phenomenon in LGS or as a core feature required for diagnosis.

Some authors include it as a criterion^5,19^ (including recent guidance from the international league against epilepsy^39^) but others do not.^4,8,21^ It is possible that children in our cohort had not yet developed tonic seizures, which may not be present at the onset of the syndrome.^20,22^ Our observations suggest the diagnosis of LGS should not rely on the presence of tonic seizures, as it would exclude children who otherwise meet the three core criteria (multiple seizure types, developmental delay, and SSW on EEG), and would miss children who may develop tonic seizures later.

### Limitations

Several limitations merit discussion. First, there were considerable missing data. Fewer than half of the children diagnosed with LGS by a physician were included, largely due to insufficient information in the medical record. Additionally, it is uncertain if the 11 children with unknown etiology have subsequently received a diagnosis, for example via more advanced genetic testing since 2014. Second, it is unclear if our cohort is representative of the general population of LGS or biased in a systematic way. We selected the cohort based on the quality of documentation in the EHR – however, “well-documented” may be a marker of the severity of illness, as these children tended to visit the healthcare system often. Our hospital-based sample is not population-based, and includes both geographic incident cases (i.e., in Bronx and Manhattan) and referred cases – this also could bias the sample towards more severely affected children. Third, we were limited by secondary use of electronic health data. Seizure frequency is often not documented in clinical notes,^40^ and parent reports of seizure frequency may underestimate seizure burden.^41^ Fourth, raw EEG tracings were not available for review, and thus our ascertainment of SSW was based on the language in the EEG reports. Further, the inter-rater reliability to describe SSW is unknown. Fifth, we did not require paroxysmal fast activity to be documented in the EEG report.

## CONCLUSIONS & NEXT STEPS

Our retrospective chart review study of children with LGS at five centers in New York City identified two opportunities to improve care for children with early-life epilepsy. First, SSW was identified as a possible biomarker of LGS as it was present early and during periods of seizure freedom. The predictive value of SSW as a biomarker would benefit from being clarified in future studies. The idea to treat early-life SSW aggressively, even during periods of seizure freedom, merits further study, potentially in a clinical trial. Second, we identified gaps in delivering first-line medication to children with IESS. Improving adherence to recommended therapy for infantile spasms would require a health delivery intervention, such as quality improvement initiatives.

## Data Availability

All data produced in the present study are available upon reasonable request to the authors and produced in the present work are contained in the manuscript.

## Acknowledgements

We are deeply grateful to Niu Tian, MD PhD and Dale Hessdorfer, PhD for their invaluable support of this project.

